# Immunopeptidomics of cutaneous leishmaniasis patients reveals the natural antigenic landscape

**DOI:** 10.64898/2026.01.06.26343515

**Authors:** Nicky de Vrij, Elise Pepermans, Louise Laurijssen, Thao-Thy Pham, Lauren Thijs, Kurt Boonen, Kadrie Ramadan, Ilse Maes, Yetemwork Aleka, Mekibib Kassa, Tigist Mekonnen, Mezgebu Silamsaw Asres, Mikias Woldetensay, Seid Hassen, Fentaw Bialfew, Feleke Tilahun, Seid Getahun Abdela, Malgorzata Anna Domagalska, Bart Cuypers, Saskia van Henten, Johan van Griensven, Pieter Meysman, Geert Baggerman, Kris Laukens, Wim Adriaensen

## Abstract

Cutaneous leishmaniasis (CL) is a skin disease caused by *Leishmania* infection, for which no licensed human vaccine exists. Protective immunity is largely T cell–mediated and depends on antigen presentation by MHC molecules, yet the naturally presented epitopes during human disease remain poorly defined. To address this gap, we performed mass spectrometry–based immunopeptidomics on lesional biopsies from 27 Ethiopian CL patients spanning the full clinical spectrum. We newly identified 333 MHC-I and 247 MHC-II epitopes from 398 *L. aethiopica* proteins, including 19 peptides and 51 antigens recurrently presented across patients of which several were also epitope-rich. Some peptides were also detected during early infection in a *L. aethiopica*–infected THP-1 monocyte model, highlighting their relevance from disease onset. Notably, despite the broad predictive coverage of NetMHCpan and NetMHCIIpan, these tools missed 20–70% of the naturally presented epitopes while predicting millions of candidates, underscoring the limitations of prediction-only vaccine pipelines. This first comprehensive map of the *Leishmania* immunopeptidome in human disease reveals conserved and prevalent antigens that can inform rational vaccine design and deepen our understanding of protective T cell responses in leishmaniasis.

## Introduction

Cutaneous leishmaniasis (CL) is a disfiguring skin disease caused by protozoan parasites of the genus *Leishmania*. Although not fatal, healed CL lesions often cause scarring or disfigurement, which can be socially stigmatizing. Ethiopian CL features a particular and diverse spectrum of clinical manifestations, ranging from localized cutaneous leishmaniasis (LCL) presenting with smaller and often self-healing localized lesions at the site of the sand fly bite, to those with mucosal involvement (referred to as mucocutaneous leishmaniasis (MCL)), and diffuse CL (DCL), which is a chronic condition and usually presents with multiple papular or nodular lesions spread across the body. Despite being endemic in more than 90 countries, there are currently no licensed vaccines against any form of human leishmaniasis. Previous vaccine development efforts either did not progress past the preclinical stage or, for the <10 candidates who did, past phase II clinical trials ^1,2^. While the reasons for this are multifactorial, subunit vaccine design is in part hindered by a lack of robust knowledge on those *Leishmania* antigens responsible for the protective response.

*Leishmania* is an intracellular parasite that resides within the phagolysosomes of the host’s antigen-presenting cells. Based on small animal models, the protective immune response to CL has been suggested to predominantly require CD4+ T helper 1 (Th1) cell-mediated intracellular killing of the parasite. In human CL, the Th1 paradigm is not as clear with CD8+ T cells also shown to contribute ^3^. This elicited T cell-based immune response requires the antigen-presenting cells of the host to present peptide epitopes derived from *Leishmania* protein antigens, on Major Histocompatibility Complex (MHC) molecules (encoded by the Human Leukocyte Antigen (HLA) genes), as a peptide-MHC complex to T cells. Due to the parasite’s hiding location in the phagolysosome, this antigen processing and presentation mechanism is targeted by the parasite as a means for immune escape. Yet, to date, we do not know how the antigenic repertoire gets skewed, and which antigens get MHC-presented in humans during natural infection and disease. In fact, only 23 *L. donovani* epitopes derived from 12 antigens (of the circa 8000 *Leishmania* proteins) have been experimentally validated to be MHC-presented and listed on the Immune Epitope Database (IEDB) (accessed on 03/09/2025). However, these mainly originated from donor-derived monocyte-derived dendritic cells that were pulsed with *L. donovani* promastigote lysate. This identifies naturally processed epitopes from crude soluble antigens rather than those derived from infection with whole live parasite. The remaining epitopes were identified from *in vitro L. major* infected bone marrow-derived dendritic cells isolated from mice, not mimicking the natural infection setting ^4,5^.

While antigen discovery for leishmaniasis vaccines was traditionally centred around low-throughput screenings of selected proteins for T cell immunogenicity, advances in computational prediction algorithms allowed high-throughput *in silico* T cell epitope screenings in leishmaniasis ^6–8^. These reverse vaccinology tools accurately predict MHC presentation of epitopes in a number of settings, such as viral infections or cancer ^9–11^. However, these tools do not take into account parasite protein abundance nor antigen availability to the antigen processing machinery, and as a consequence, may ultimately fail to identify clinically relevant MHC-presented T cell epitopes for *Leishmania*. Recent advancements in mass spectrometry-based immunopeptidomics have dramatically increased the sensitivity and throughput capacity, enabling the profiling of endogenous antigen presentation in *in vitro* settings or directly in clinical samples within the context of intracellular pathogens, as shown for *Mycobacterium tuberculosis* and *Trypanosoma cruzi* ^12–14^.

In this study, we leveraged this powerful technique for a first-in-kind identification of 626 novel MHC-presented epitopes derived from 410 *Leishmania* proteins in a *L. aethiopica*–infected THP-1 monocyte model and lesional biopsies of Ethiopian CL patients.

## Methods

### Ethics

This study on biobanked skin samples of the SSG-Allopurinol study (clinicaltrials.gov identifier: NCT04699383, study registered on 19 October 2020) and samples of the SpatialCL study (clinicaltrials.gov identifier: NCT05332093, study registered on 8 March 2022) was approved by the Ethiopian National Research Ethics Review Committee (NRERC; 17/292/973/22), the University of Gondar Institutional Review Board (VP/RTT/05/647/2022), and the Institute of Tropical Medicine Antwerp Institutional Review Board (1539/21). All participants provided written informed consent and the study was carried out in accordance with both national and international guidelines (Helsinki declaration, Good Clinical Practices, and local regulations).

### Study design and study population

Biobanked, in liquid nitrogen flash-frozen lesional punch biopsies from the index lesions were obtained from patients enrolled in two clinical studies on Ethiopian CL, namely the SSG-Allopurinol study and the SpatialCL study using a 3mm or 4mm (Kai Medical, Solingen, Germany) punch biopsy device, respectively. In total, 27 patients across the entire Ethiopian CL spectrum were included (of which 13 (48.1%) from SpatialCL and 14 (51.9%) from SSG-Allopurinol) for use in mass spectrometry-based immunopeptidomics (**Figure 1**). Patients from the SSG-Allopurinol study will be referred to as cohort 1, while patients from the SpatialCL will be referred to as cohort 2. The patients were selected based on a combination of availability and clinical presentation to select a representative subset of the complete Ethiopian CL spectrum. Based on clinical assessment by the treating clinicians, cohort 1 consisted of 3 (21.4%) LCL patients, 10 (71.4%) MCL patients, and 1 (7.2%) DCL patient. Cohort 2 consisted of 5 (38.5%) LCL patients, 5 (38.5%) MCL patients, and 3 (23%) DCL patients. It should, however, be noted that this classification (based on Ethiopian guidelines, and borrowed from the South American setting) leads to many ambiguous cases and differential classification between dermatologists in practice ^15^.

To study epitope/antigen MHC-presentation early after infection, we studied *L. aethiopica*-infected *in vitro* THP-1 monocyte cell lines differentiated to macrophages. *L. aethiopica* was selected as it is the predominant *Leishmania* species in Ethiopia. Altogether, these immunopeptidomics experiments were performed in four batches (**Table 1**), including: the *in vitro L. aethiopica-*infected and HLA-restricted cell lines across early infection timepoints (5h and 24h post-infection for batch #1, and 72h post-infection for batch #2), and different human CL patient skin lesion biopsy samples across diverse HLA allele backgrounds ran on two different mass spectrometry machines (Batch #3 on TimsTOF-Pro (Bruker Daltonics, Bremen, Germany) and batch #4 on the more sensitive TimsTOF-SCP (Bruker Daltonics, Bremen, Germany)).

**Table 1.**
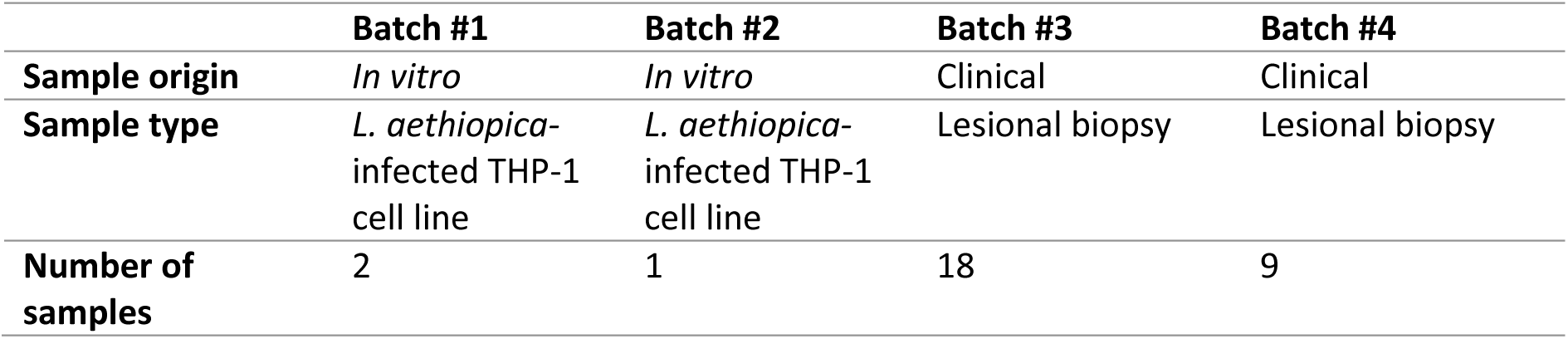
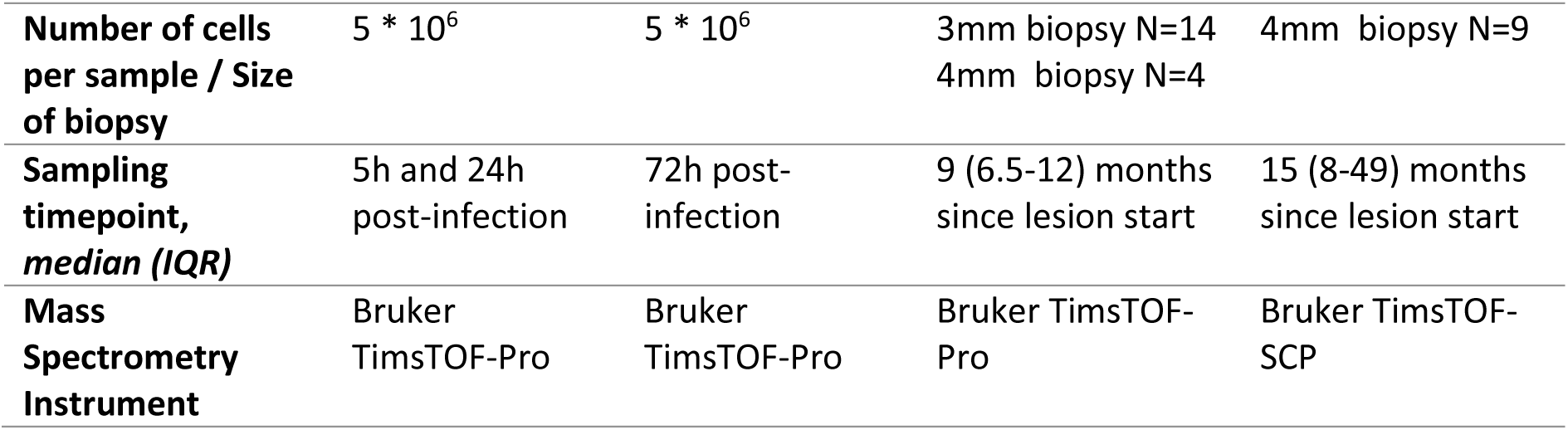
Description of the different immunopeptidomics batches with varying sample conditions and instrumentation. Clinical sampling timepoint is provided as median (IQR).

|  | Batch #1 | Batch #2 | Batch #3 | Batch #4 |
| --- | --- | --- | --- | --- |
| **Sample origin** | *In vitro* | *In vitro* | Clinical | Clinical |
| **Sample type** | *L. aethiopica*-infected THP-1 cell line | *L. aethiopica*-infected THP-1 cell line | Lesional biopsy | Lesional biopsy |
| **Number of samples** | 2 | 1 | 18 | 9 |
| Number of cells per sample / Size of biopsy | 5 * 10<sup>6</sup> | 5 * 10<sup>6</sup> | 3mm biopsy N=14<br>4mm biopsy N=4 | 4mm biopsy N=9 |
| Sampling timepoint, median (IQR) | 5h and 24h post-infection | 72h post-infection | 9 (6.5-12) months since lesion start | 15 (8-49) months since lesion start |
| Mass Spectrometry Instrument | Bruker TimsTOF-Pro | Bruker TimsTOF-Pro | Bruker TimsTOF-Pro | Bruker TimsTOF-SCP |

**Figure 1.**
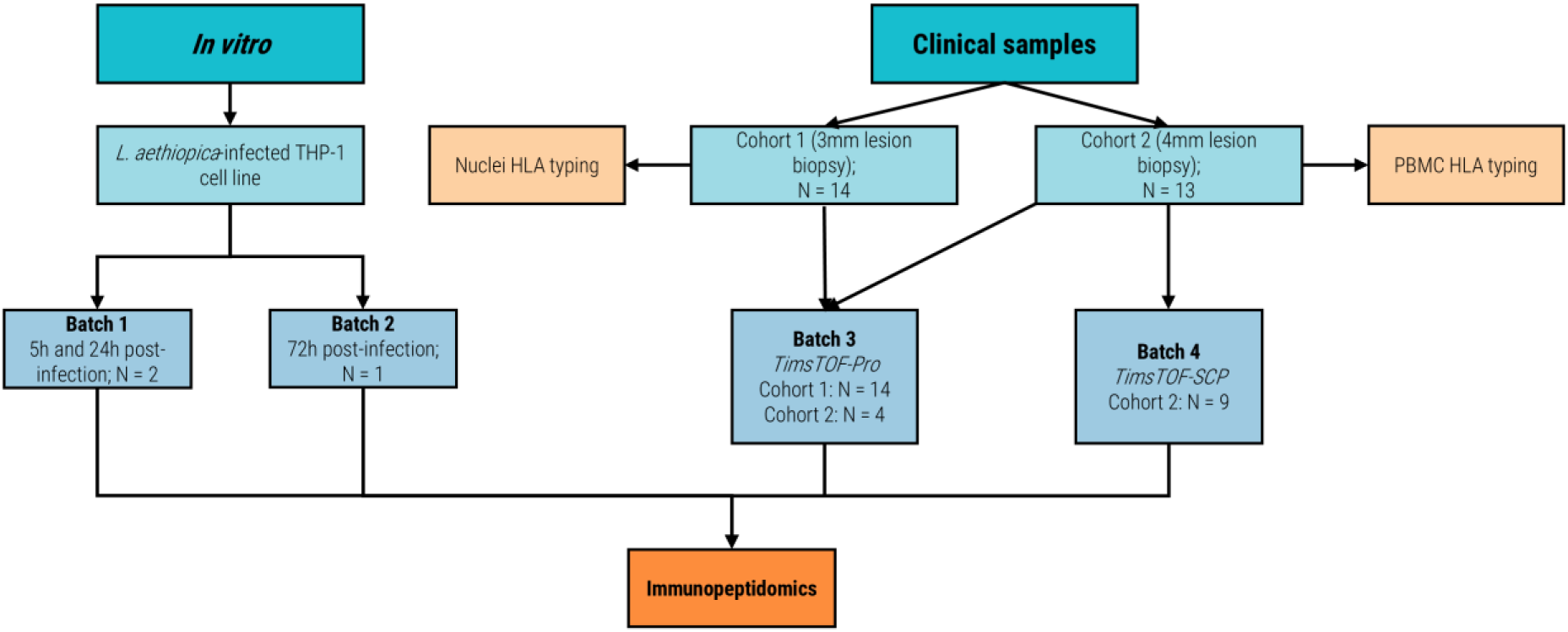
Flowchart of study design. This flowchart describes the study design, number of samples in each batch, from which study they originated, and chosen methodology. N: number of samples

### THP-1 cell culture and *in vitro* infection with *L. aethiopica*

THP-1 monocytes (ATCC TIB-202) were cultured in T25, T75, and T175 culture flasks (Greiner Bio-one, Belgium) using a Roswell Park Memorial Institute (RPMI) (Biowest, U.S.A.) medium supplemented with 10% Fetal Calf Serum (Biowest, U.S.A.) and 50µM 2-mercapto-ethanol (Sigma-Aldrich, Belgium). These THP-1 monocytes were cultured until 5 million cells could be obtained. Next, the THP-1 monocytes were differentiated to macrophages by overnight stimulation with 0.1 µM Phorbol 12-Myristate Acetate (PMA, Thermo Fisher, Belgium). After differentiation to macrophages, the cells were infected with *L. aethiopica* promastigotes (MHOM/ET/72/L100, passage Rx+29). Infected macrophages were harvested at either 5 hours (batch 1), 24 hours (batch 1), or 72 hours (batch 2) post-infection for use in mass spectrometry-based immunopeptidomics.

### Mass spectrometry-based immunopeptidomics

The *L. aethiopica-*infected THP-1 macrophages and 18 CL patient lesion biopsy samples (batch 3) were processed by the Centre for Proteomics of the University of Antwerp. The clinical samples were homogenized by sequential homogenization using a Next Advance BulletBlender Gold bead homogenizer, in a lysisbuffer containing 150mM NaCl, 50mM tris hydroxymethylaminomethane at pH 8, one tablet of cOmplete™ ethylenediaminetetraacetic acid (EDTA)-free anti-protease (Roche, Germany) per 50mL, and 0.1% Fos-choline-14 diluted ULC/MS-grade water, whereas the *L. aethiopica*-infected THP-1 macrophages were lysed using the same buffer without bead homogenization. The samples were then incubated for 1 hour at 4°C. Next, the samples underwent several centrifugation rounds at 4°C to pellet down the nuclei (see HLA genotyping section), and to remove unsolubilized membranes, purifying the MHC complexes. Next, lysates were pre-cleared on empty protein A/G-sepharose bead columns to remove non-specific binders. After this, MHC class I and class II complexes were subjected to sequential affinity purification by binding of the pre-cleared lysate to MHC immuno-affinity columns. These columns were generated by cross-liking protein A sepharose beads to anti-HLA I antibody (clone W6/32) and anti-HLA II antibodies (clones L243, 1a3, and B7/21). After batch binding, beads were sequentially washed with a series of wash buffers to non-specifically bound material and residual salt and detergent, followed by a mild acid elution with 10% formic acid to elute MHC complexes from the column and dissociate immunopeptides. Subsequently, immune peptides were separated from the larger components of the MHC complexes by C18 solid phase extraction. Next, the peptides were loaded for measurement on a TimsTOF-Pro (Bruker Daltonics, Bremen, Germany) connected to an Evosep One chromatography system (Evosep Biosystems, Denmark). The Evosep was run with a 44 minutes default gradient and an EV1106 column. The Parallel Accumulation–Serial Fragmentation (PASEF) DDA method without pre-defined polygone was employed to select precursor ions for fragmentation involving 1 TIMS-MS scan and 10 PASEF MS/MS scans. The TIMS-MS survey scan spanned 0.70 to 1.40 Vs/cm2 and 100–1,700 m/z, with a ramp time of 166ms for MHC-I peptides, and 100ms for MHC-II peptides, while a linear collision energy ramp from 20 to 59 EV with an ion mobility range from 1.6 to 0.6 Vs/cm2 was used for peptide fragmentation.

Lesion biopsy samples from 9 CL patients (batch 4) were processed by ImmuneSpec (Niel, Belgium), using a more sensitive workflow and more sensitive mass spectrometry device. This workflow is similar to the above, with the following steps adapted: these samples were homogenized in ImmuneSpec’s propietary lysisbuffer by sequential homogenization steps using a Precellys bead homogenizer (Bertin Technologies, France) followed by 1 hour incubation (lysis). Lysates were precleared on empty protein A-sepharose beads (to remove non-specific binders) prior to binding on anti-HLA I-Sepharose beads (antibody clone W6/32 crosslinked to protein A Sepharose), and sequential binding on anti-HLA-II-Sepharose beads (antibody clones L243, 1a3, B7/21 crosslinked to protein A Sepharose). After C18 solid phase extraction, the eluted peptides were dried, resuspended in Mobile Phase A, and loaded on evotips prior to measurement on a Bruker Daltonics TimsTOF-SCP coupled to an Evosep One Nano-LC.

PEAKS Online 11 (Bioinformatics Solutions Inc., Canada) was used for MS data analysis. A database search was performed against the reviewed Uniprot Human Proteome (Swiss-Prot only) database (release 2024_03, 42.490 entries, downloaded March 2024), concatenated with the reference proteome of *L. aethiopica* strain L147 (version 67; 8.533 entries) acquired from the TriTryDB ^16^. Error tolerance was set at 20 ppm and 0.05 Da for precursor ions and fragmentation ions, respectively. Digestion was set as unspecific and no enzyme was specified. Oxidation (M) and Deamidation (NQ) were selected as variable modifications. A false discovery rate (FDR) of 1% was set at peptide level.

### HLA genotyping using NanoTYPE™

DNA was extracted from either patient whole blood collected in 10mL Lithium-Heparine tubes (BD Vacutainer, Erembodegem, Belgium; Cohort 2 study samples) or the nuclei byproducts of the mass spectrometry-based immunopeptidomics assay (Cohort 1 study samples) (Figure 1), using the Maxwell® RSC 48 instrument (Promega, Wisconsin, U.S.A.) with the Maxwell® RSC Whole Blood DNA kit (Promega, Wisconsin, U.S.A.) according to the manufacturer’s instructions. Next, the concentration of the DNA was measured using the Qubit 1X dsDNA BR assay (ThermoFisher, Waltham, U.S.A.) on a Qubit Fluorometer 4.0 device (ThermoFisher, Waltham, U.S.A.). A total of 200ng of participant’s DNA was used for HLA genotyping using the Oxford Nanopore Technologies (ONT)-sequencing-based NanoTYPE™ assay (Omixon, Budapest, Hungary) according to the manufacturer’s instructions. In brief, an enrichment PCR was performed using the HLA Multi Primer Mix and reagents included in the NanoTYPE™ kit. Next, amplicons were quantified using the Qubit 1X dsDNA BR assay (ThermoFisher, Waltham, U.S.A.) on a Qubit Fluorometer 4.0 device (ThermoFisher, Waltham, U.S.A.), and a total of 200ng of amplicon was transferred to a new tube. A barcoding step was then performed using the Rapid Barcoding Plate provided in the Rapid Barcoding 96 Kit (SQK-RBK110.96; ONT, Oxford, U.K.). After this, samples were pooled (between 8-12 samples per library), adding 8.5 µl of barcoded amplicons per sample to a library. Next, resulting pooled libraries were subjected to size selection and purification using the AMPure XP beads (Beckman Coulter, Brea, U.S.A.) also provided in the Rapid Barcoding 96 Kit. A total of 10 µl of purified library was then transferred to a new tube, and 1 µl of Rapid Adapter F (provided in the Rapid Barcoding 96 Kit) was added to the library. The resulting library with added adapters was then mixed with 37.5 µl of Sequencing Buffer II and 25.5 µl of Loading Beads II to load on to a R9.4.1 Flow Cell (ONT, Oxford, U.K.) for subsequent sequencing on a MinION Mk1B instrument (ONT, Oxford, U.K.) with MinKNOW version 22.05.5. Libraries were sequenced for at least 1 hour per sample in a library. The resulting FAST5 files were basecalled using Guppy version 6.1.5 using the high-accuracy model. Finally, basecalled FASTQ files were used for HLA genotype inference with the NanoTYPER v2.0.0 software (Omixon, Budapest, Hungary).

### Immunopeptidomics data analysis

Peptides were assigned MHC restrictions using the MHCMotifDecon 1.2 software with default settings using the HLA genotyping assay data ^17^. All other downstream analyses on the PEAKS Online 11 output was performed in R version 4.3.3. *Leishmania* MHC-presented peptides were defined as those mapping exclusively to the *Leishmania* proteome, and using MHC class-specific length thresholds of 8-14mer and 12-21mer peptides for MHC-I and MHC-II, respectively ^17^. To verify if our observed *L. aethiopica* peptides are similar in sequence to prior published *Leishmania* epitopes, we queried the IEDB and calculated Levenshtein distances using the *stringdist* package version 0.9.1. Next, heatmaps of antigen or peptide overlap between patients were created using the *complexHeatmap* package version 2.18.0. Next, we calculated the summed number of MHC-presented peptides derived from an antigen, which can shape the immunodominance of an antigen ^12^.

### Gene Ontology Enrichment

A gene ontology (GO) enrichment test was performed on all the MHC-presented *Leishmania* antigens using the enricher function (default settings) of the clusterProfiler package at version 4.10.1 in R version 4.3.3 with curated *L. aethiopica* gene ontologies acquired from the TriTrypDB ^16,18^. This was performed on all *Leishmania* antigens, and separately for MHC-I and MHC-II. Only Benjamini-Hochberg-adjusted p-values below 0.05 were considered significant.

### MHC epitope-binding predictions

Proteome-wide prediction of *L. aethiopica* epitopes was performed using the reference proteome of *L. aethiopica* strain L147 (version 67), acquired from the TriTrypDB, as input for NetMHCpan 4.1 and NetMHCIIpan 4.3 ^9,10,16^. This was performed for the HLA genotypes of the THP-1 monocyte (ATCC TIB-202) cell line. The NetMHCpan peptide lengths were set to 8-14mer, which deviates from the default range of fragment lengths (between 8-11mer), and the NetMHCIIpan peptide lengths were set to 12-21. These fragment lengths were chosen to match the length thresholds applied in the immunopeptidomics data analysis. Subsequently, only predicted strong and weak binders, according to the default %Rank cut-offs, were included for further analysis.

## Results

### Patient characteristics

Our study included a total of 27 patients, of which 23 (85%) were male with a median age of 22 years, matching prior observations that CL in Ethiopia affects mostly young adults (Table 2) ^15^. The CL episodes persisted for a median of 9 (6.5-15) months prior to sampling, and presented with a median of 1 (1-2) lesion of approximately 6 (4-7) cm predominantly located on the face (78%). Of the 27 CL patients, 4 (14.8%) had more than 5 lesions.

**Table 2.** CL patient characteristics. Continuous variables were represented as medians with interquartile ranges (IQR) and categorical data as numbers and proportions.

|  | Total (n=27) | Cohort 1 (n=14) | Cohort 2 (n=13) |
| --- | --- | --- | --- |
| **Socio-demographic characteristics** |  |  |  |
| Age in years, median (IQR) | 22 (18-33) | 25.5 (19-39) | 20 (18-26) |
| Male, n (%) | 23 (85) | 12 (85.7) | 11 (84.6) |
| **Clinical history and lesion characteristics** |  |  |  |
| Current CL episode duration in months, median (IQR) | 9 (6.5-15) | 9 (7-12) | 8 (6-16) |
| Number of lesions, median (IQR) | 1 (1-2) | 1 (1-2) | 1 (1-3) |
| Size (largest diameter), median (IQR) | 6 (4-7) | 6 (6-8) | 4 (3-6) |
| **Location of lesion biopsy was taken from, number (%)** |  |  |  |
| Face | 21 (78) | 12 (85.7) | 9 (69.2) |
| Limbs | 3 (11) | 1 (7.15) | 2 (15.4) |
| Other | 3 (11) | 1 (7.15) | 2 (15.4) |
| **Index lesion presentation. number (%)** |  |  |  |
| Plaque | 18 (66.7) | 14 (100) | 4 (30.8) |
| Scaly | 19 (70.4) | 12 (85.7) | 7 (53.8) |
| Crusted | 17 (63.0) | 7 (50) | 10 (76.9) |
| Papular | 7 (25.9) | 6 (42.9) | 1 (7.7) |
| Erythematous | 13 (48.1) | 3 (21.4) | 10 (76.9) |
| Hyperpigmented | 9 (33.3) | 9 (64.3) | 0 (0) |
| Swollen | 12 (44.4) | 2 (14.3) | 10 (76.9) |
| Nodular | 11 (40.7) | 5 (35.7) | 6 (46.2) |
| Ulcerated | 1 (3.7) | 1 (7.1) | 0 (0) |

### Mass spectrometry-based identification of *L. aethiopica* peptides presented by MHC

To identify MHC-presented *Leishmania* antigens and epitopes, we performed mass spectrometry-based immunopeptidomics across different experiments and conditions (see Figure 1).

We identified a total of 85.920 and 74.804 different unique MHC-I and MHC-II peptides, respectively **Fig. S1A)**. Of these, 7.231 MHC-I and 3.452 MHC-II peptides were identified in the *in vitro* samples of batch #1 and batch #2, and 78.689 MHC-I and 21.003 MHC-II peptides were identified in the clinical samples of batch #3 and batch #4. The eluted peptide length distributions summed across experiments **(Fig. S1B)**, or within each experiment **(Fig S2)**, were consistent with expected MHC-I and MHC-II peptide length distributions. Peptides of 9mer length were the predominant eluted peptide length for MHC-I, and MHC-II showed a peak around 15mer peptides ^19,20^.

In total, 511 MHC-I and 411 MHC-II presented peptides mapped exclusively to *L. aethiopica* (**Fig. 2A**). As expected, a higher yield was reached in batch #4 compared to batch #3 due to the use of a more sensitive mass spectrometer. Further selection of high-confidence MHC-I peptides with a length restriction between 8-14mer and MHC-II peptides between 12-21mers (dashed lines on **Fig. 2B, Fig. S1B, and Fig. S2-3**) resulted in 379 MHC-I and 271 MHC-II *L. aethiopica* peptides across 443 *L. aethiopica* antigens included for further analyses **(listed in Supplementary Excel)**. Of these, 333 MHC-I and 247 MHC-II *Leishmania* peptides across 398 *Leishmania* antigens were identified in clinical samples. Of note, 245 (37.7%) of the 650 MHC-presented *Leishmania* peptides were identified from a single DCL patient. None of these T cell epitopes were previously reported on the IEDB, and all identified peptides deviated by at least a Levenshtein distance of 4 (being largely dissimilar) from those epitopes previously described ^4^.

**Figure 2.**
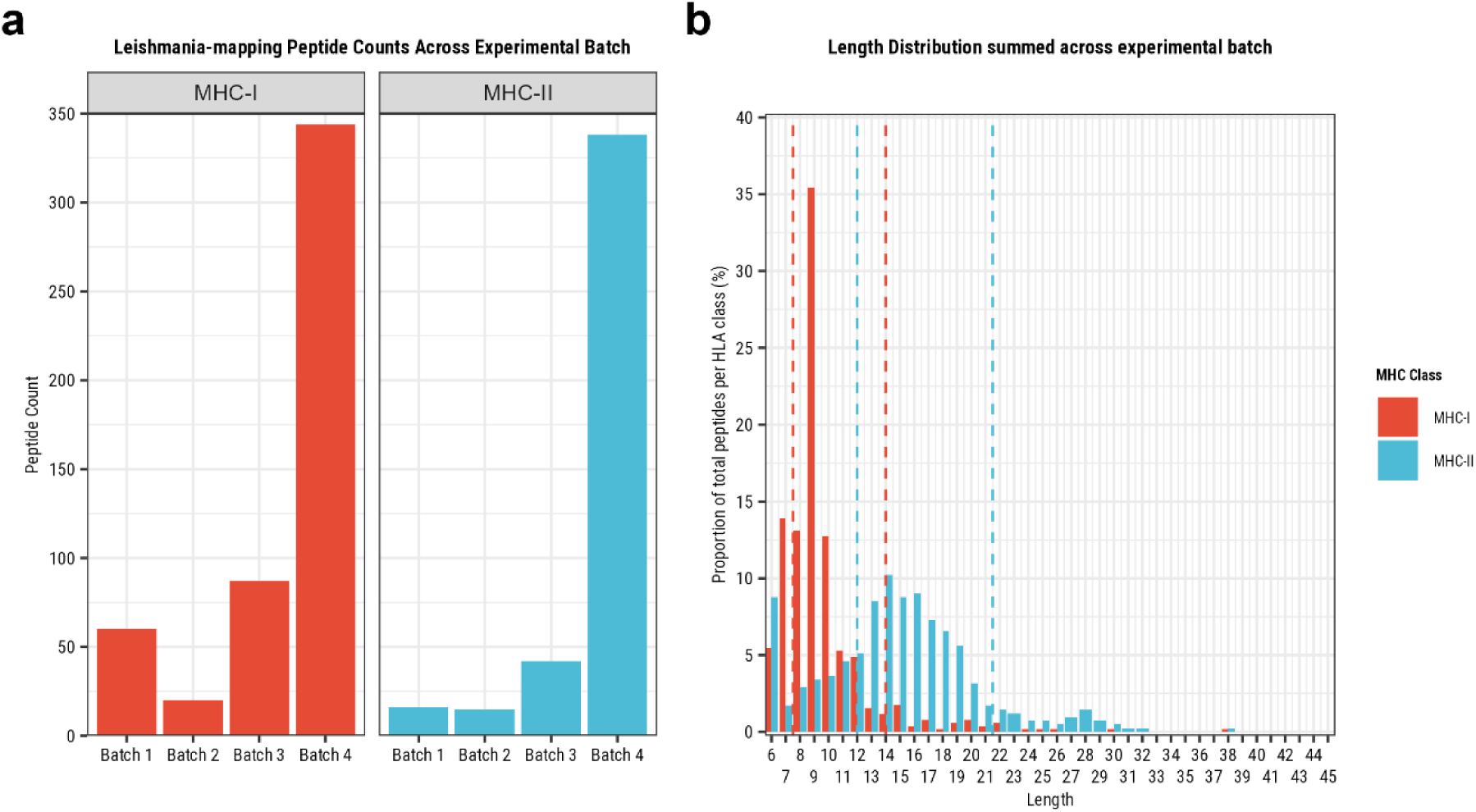
MHC-presented peptide counts and lengths of peptides exclusively mapping to L. aethiopica proteins, and the lengths of these peptides across experiments. Batch #1 and batch #2 are in vitro samples, while batch #3 and batch #4 are from clinical samples (See methods). (A) The peptide count per experiment by MHC class, in red for MHC-I and blue for MHC-II. (B) The length distribution of all MHC-presented peptides by MHC class, in red for MHC-I and blue for MHC-II. The red dashed lines represent the 8-14mer length threshold for selecting higher-confidence MHC-I peptides, and the blue dashed lines represent the 12-21mer length threshold for selecting higher-confidence MHC-II peptides.

### Shared epitope and antigen presentation across patients

Next, we studied whether there was any overlap in epitope or antigen presentation between patients with different timings from infection, and with early *in vitro* infected samples. Those epitopes or antigens that are shared between individuals represent potential immunoprevalent targets, a measure for how prevalent an eventual immune response would be in a given population, which correlates with the immunodominance of a response ^21–24^.

Despite the HLA diversity between patients, and mass spectrometry detection limits revealing only a subset of all presented epitopes, 19 exact peptides (13 MHC-I, 6 MHC-II) were shared between two patients (**Fig. 3**). In one case, a peptide was shared between two long-term CL patients and was also already presented as early as 24h post-infection in a *L. aethiopica*-infected THP-1 cell line, namely *IINEPTAAAIAYGLN[Deamidated]K*, derived from the antigen Heat-shock Protein (Hsp70; LAEL147_000494700). Six MHC-presented peptides, shared between two patients, were derived from antigens annotated as ‘hypothetical protein’. Patients sharing an exact peptide had at least one or more matching HLA allele with the other patient, except in the case of the peptides *AAPAGAAAL* and *ASASASAAL* where patients only carried the closely related HLA-C*07:01 and HLA-C*07:02, and C*03:03 and C*03:04 alleles, respectively, which likely share peptide specificity **(Supplementary Excel 2)**. Due to limited HLA genotyping quality, HLA matches could not be determined and confirmed for all. For the peptide *YLTAELLEL*, no HLA genotype match or close match could be observed, suggesting that there is likely overlap in the binding peptide repertoires of two HLA alleles that are not closely related. Two peptides, *M[Oxidation]PPQFGSGRPF* and *RLLDVVYNA*, were presented in both the 5h and 24h post-infection in the *L. aethiopica*-infected THP-1 cell line samples, showing good concordance between samples of the same batch. The peptide *IKSGTAHKSFVD* was present at both 5h and 72h post-infection in these *in vitro* samples, also suggesting the detection of peptides is robust across batches.

**Figure 3.**
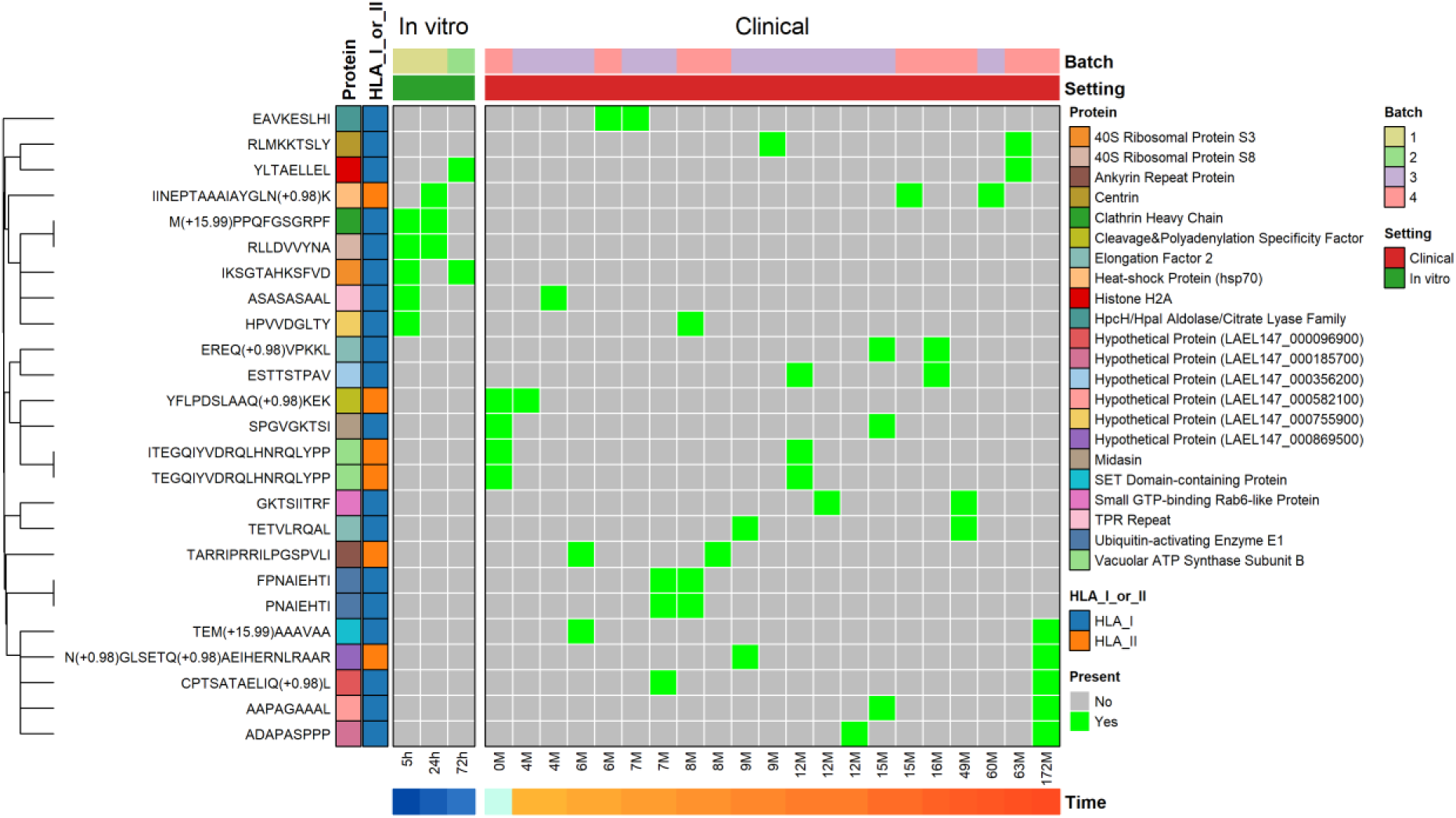
Exact MHC-presented epitope matches between samples. Every column represents a sample and every row indicates an MHC-presented peptide. On the top of the plot, the experimental batch, and whether it was an in vitro or clinical sample, are depicted. On the bottom, the time post-infection (for in vitro samples) or current CL lesion duration (for clinical samples) is shown. Rows were clustered to identify groups of epitopes presented in the same samples.

As exact peptides might not be shared but still derived from the same protein, we subsequently analysed which proteins were shared across patients. Here, 51 antigens were presented in a minimum of two patients and a maximum of 5 patients (**Fig. 4**). Of these, 43 (84.3%) were shared across two patients, 7 (13.7%) were shared across three patients, and one (2%, Elongation Factor 2 (EF-2; LAEL147_0000067)) was shared between five patients (**Fig. 4**). Of the five patients sharing EF-2 presentation, four had two matching HLA alleles, namely HLA-B*49:01, and HLA-C*07:01, while the other patient had the closely related HLA-C*07:02 allele.

When including the *in vitro* samples, 66 (14.9% of total antigens) antigens were shared between a minimum of two samples and maximum of 5 samples (**Fig. 4**). Of these 66 shared antigens, 54 (81.8%) were shared between two samples, 6 (9.1%) were shared between three samples, 4 (6.1%) were shared between four samples, and 2 (3%, namely EF-2 and Hsp70) were shared between five samples. Three antigens, Clathrin heavy chain (LAEL147_000825100) and two copy number variants of 40S Ribosomal Protein S8 (LAEL147_000396600; LAEL147_000396700), were presented in the 5h and 24h post-infection *in vitro* samples, showing good concordance between samples of the same batch. Two copies of 40S Ribosomal Protein S3 were presented in the 5h and 72h post-infection *in vitro* samples, and a copy of Heat-shock Protein Hsp70 and Fusaric Acid Resistance Protein (LAEL147_000168900) were presented in the 24h and 72h post-infection *in vitro* samples, showing good concordance between all *in vitro* samples. As expected, the DCL patient where we identified the most MHC-presented peptides from also had the highest number of antigens shared between other patients with 27 (40.9%) of 66 shared antigens. Of the 66 antigens, 19 (24.2%) were annotated as ‘hypothetical protein’, for which there is no known function yet.

**Figure 4.**
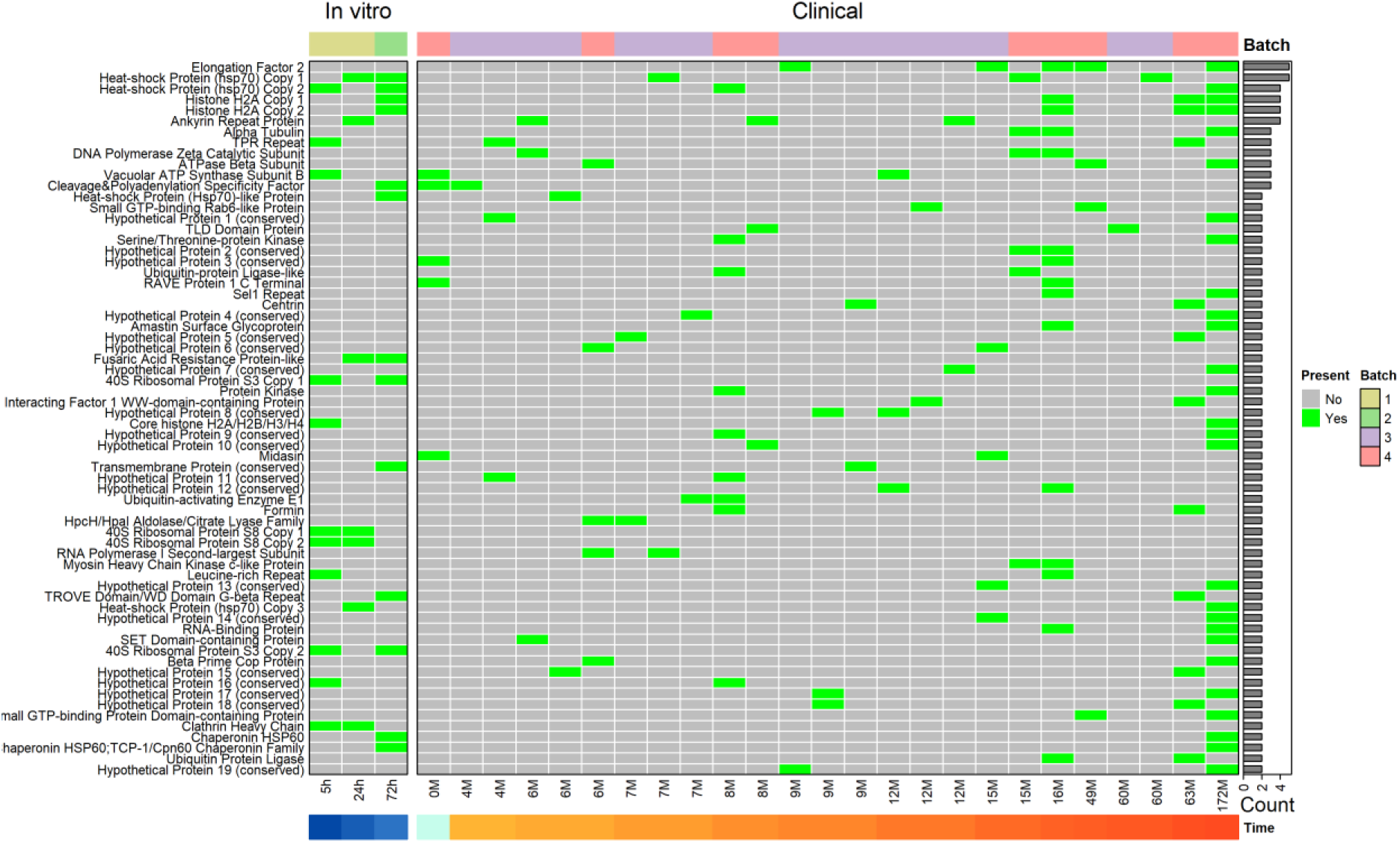
MHC-presented L. aethiopica antigen matches across samples. Every column represents a sample and every row indicates an MHC-presented antigen. An antigen is considered present in a sample if at least one MHC-presented Leishmania peptide was derived from it. On the top of the plot, the experimental batch, and whether it was an in vitro or clinical sample, are depicted. On the bottom, the time post-infection (for in vitro samples) or current CL lesion duration (for clinical samples) is shown. On the right, the total number of samples an antigen is present in is shown. Rows were clustered to identify groups of antigens presented in the same samples.

### Identifying epitope-rich antigens

As immunodominance can be reflected by a high number of T cell epitopes across the protein (epitope-richness), we next looked at the number of MHC-presented peptides per identified *L. aethiopica* antigen.

EF-2, Alpha Tubulin (LAEL147_000015900), and glyceraldehyde 3-phosphate dehydrogenase (GAPDH; LAEL147_000578000) had 5 MHC-presented peptides across 5, 3, and 1, patients, respectively (**Fig. 5A**). Next, Histone H2A (LAEL147_000533800; LAEL147_000533900) and 10kDa Heat Shock Protein (LAEL147_000431900) had 4 MHC-presented peptides across 3 and 1 patients, respectively. DNA Polymerase Zeta Catalytic Subunit (LAEL147_000368800), and ATPase Beta Subunit (LAEL147_000412300), Hsp70, Ankyrin Repeat Protein (LAEL147_000770300), all had 3 peptides across 3 patients each. Myosin Heavy Chain Kinase C-like Protein (LAEL147_000406700) also had 3 MHC-presented peptides but only in 2 patients.

When including the *in vitro* samples of early infection, the antigen with the highest number of eluted peptides was Histone H2A with 6 peptides across four samples (**Fig. 5B**). In addition, the one peptide of this antigen that was shared between a patient and an *in vitro* sample was already presented as early as 72h post-infection (**Fig. 3**). The antigens with the second highest number of eluted peptides were EF-2, Hsp70, Alpha Tubulin, 40S Ribosomal Protein S33 (LAEL147_000442300), and GAPDH with 5 epitopes per antigen (**Fig. 5B**). While peptides of EF-2 and Hsp70 were presented in 5 different samples, and Alpha Tubulin in 3 samples, peptides of 40S Ribosomal Protein S33 and GAPDH were only presented in a single sample. A second gene copy number variant of Hsp70 (LAEL147_000511600) and Ankyrin Repeat Protein had 4 peptides presented in four samples, and 10kDa Heat Shock Protein also had 4 peptides but presented only in a single sample.

**Figure 5.**
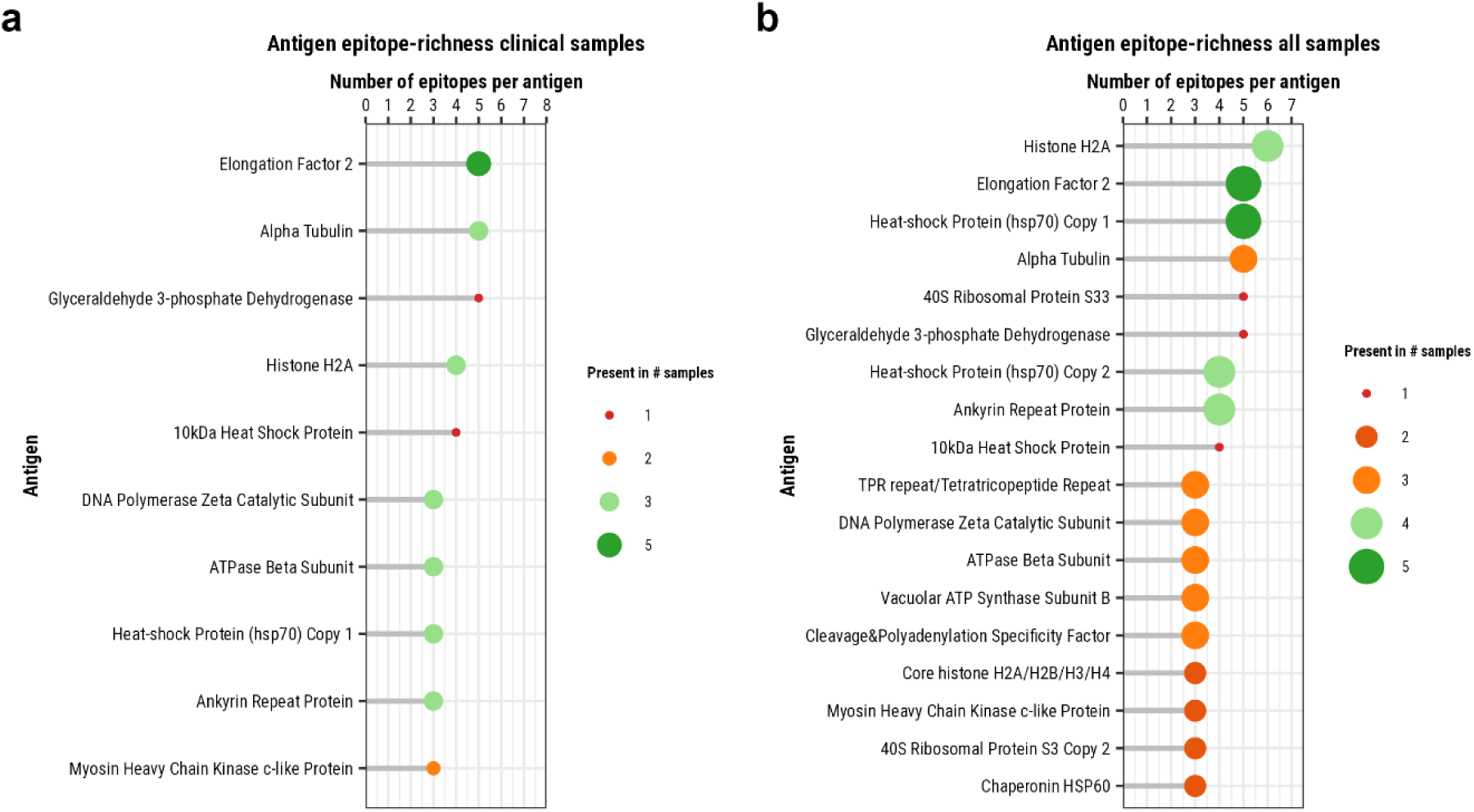
Antigen epitope-richness. The number of identified MHC-presented high-confidence L. aethiopica peptides per MHC-presented antigen, summed up across patients (a) or all samples (b). Size and colour indicate in how many samples or patients a protein is presented.

### Biological characteristics and conservation between species of the MHC-presented *L. aethiopica* antigens

To further characterize the biological characteristics of the naturally presented *L. aethiopica* immunopeptidome, the cellular location and biological function of the MHC-presented *Leishmania* antigens was analysed.

We first performed a GO enrichment test on all the MHC-presented *Leishmania* antigens, and only the terms “Ribosome”, “Structural constituent of ribosome”, and “Translation”, were significantly over-represented **(Fig. S4)**. Performing this GO over-representation test for only MHC-I-presented or MHC-II-presented *Leishmania* antigens resulted in the terms “Nucleosome”, “DNA binding”, “Ribosome”, “Structural constituent of ribosome”, and “ATPase-coupled transmembrane transporter activity” for MHC-I, and “Ribosome”, “Structural constituent of ribosome”, and “Translation” for MHC-II. This suggests most of the observed MHC-presented *Leishmania* antigens tend to be involved in mRNA to protein translation as part of ribosomal machinery. Although for MHC-I in particular, nucleosomal and transmembrane proteins with DNA-binding, and ATPase activity, tend to be overrepresented as well.

All antigens have orthologues in four other common *Leishmania* species (*L. major*, *L. donovani*, *L. infantum*, and *L. braziliensis)*, and share high degrees of sequence identity (68.95%-100%, **Table 3**), indicating a high level of protein conservation across species.

**Table 3.** Biological description of the top 10 antigens that are shared across multiple (>=3) patients. . Descriptions include the protein length, the Gene Ontology identifier and terms, and the sequence identity between L. aethiopica and the four most common orthologous species of the Leishmania genus, based on NCBI’s BLAST tool.

| Protein (Accession) | Protein Length | GO Terms | Orthologous species (Sequence identity %) |
| --- | --- | --- | --- |
| Elongation Factor 2 (LAEL147_000006700) | 487 | GTPase Activity<br>GTP Binding | *L. major* (99.59%), *L. donovani* (99.59%), *L. infantum* (99.59%), *L. braziliensis* (99.59%) |
| Histone H2A (LAEL147_000533800; LAEL147_000533900) | 132 | Nucleosome assembly<br>DNA binding<br>Nucleosome<br>Nucleus | *L. major* (95.45%), *L. donovani* (96.97%), *L. infantum* (97.73%), *L. braziliensis* (79.65%) |
| Alpha Tubulin (LAEL147_000015900) | 370 | NA | *L. major* (100%), *L. donovani* (99.73%), *L. infantum* (100%), *L. braziliensis* (99.73%) |
| Tetratricopeptide Repeat Protein (LAEL147_000169100) | 1079 | NA | *L. major* (95.32%), *L. donovani* (91.83%), *L. infantum* (91.44%), *L. braziliensis* (80.3%) |
| DNA Polymerase Zeta Catalytic Subunit (LAEL147_000368800) | 3094 | Nucleobase-containing<br>Compound Metabolic Process<br>DNA Replication<br>Nucleotide/Nucleic Acid/DNA Binding<br>DNA-directed DNA Polymerase Activity | *L. major* (92.41%), *L. donovani* (90.86%), *L. infantum* (90.63%), *L. braziliensis* (68.95%) |
| ATPase Beta Subunit (LAEL147_000412300) | 525 | Proton motive force-driven ATP Synthesis<br>ATP metabolic process<br>Proton transmembrane transport<br>Nucleotide binding<br>ATP binding<br>Ribonucleoside Triphosphate phosphatase activity<br>ATPase-coupled transmembrane transporter activity<br>Proton-transporting ATP synthase activity<br>Proton-transporting two-sector ATPase complex | *L. major* (99.81%), *L. donovani* (99.38%), *L. infantum* (99.81%), *L. braziliensis* (97.52%) |
| Heat-Shock Protein Hsp70 (LAEL147_000494700; LAEL147_000511600) | 659; 259 | NA | *L. major* (96.05%), *L. donovani* (95.9%), *L. infantum* (95.75%), *L. braziliensis* (91.5%) |
| Vacuolar ATP Synthase Subunit B (LAEL147_000508100) | 495 | ATP metabolic process<br>Proton transmembrane transport<br>ATP binding<br>ATPase-coupled transmembrane transporter<br>Proton-transporting two-sector ATPase complex<br>Proton-transporting V-type ATPase V1-domain | *L. major* (99.6%), *L. donovani* (99.19%), *L. infantum* (99.19%), *L. braziliensis* (95.35%) |
| Cleavage and Polyadenylation Specificity Factor-like Protein (LAEL147_000635500) | 1541 | Nucleid acid binding<br>Nucleus | *L. major* (97.14%), *L. donovani* (96.82%), *L. infantum* (96.82%), *L. braziliensis* (85.62%) |
| Ankyrin Repeat Protein (LAEL147_000770300) | 3774 | NA | *L. major* (95.63%), *L. donovani* (94.06%), *L. infantum* (94.06%), *L. braziliensis* (81.8%) |

### Overlap between MS-based immunopeptidomics and peptide-MHC binding prediction tools

As a final step, we wanted to evaluate the overlap between our experimentally acquired results with the most widely used *in silico* peptide-MHC binding prediction tools (NetMHCpan and NetMHCIIpan). First, we compared *L. aethiopica* proteome-wide prediction outputs to the experimentally acquired immunopeptidomics data of the HLA-restricted *L. aethiopica*-infected THP-1 cell line (**Fig. 6A and Fig. 6B**). In total, NetMHCpan and NetMHCIIpan predicted 1.587.865 MHC-I and 6.989.036 MHC-II peptides, respectively. For MHC-I, NetMHCpan correctly predicted 15 (36.6%) of the 41 epitopes observed to be MHC-presented in the *in vitro* immunopeptidomics experiments (**Fig. 6A**). Similarly, NetMHCIIpan predicted 7 (29.2%) of the 24 MHC-II epitopes observed in the *in vitro* immunopeptidomics experiments (**Fig. 6B**). Next, we compared *L. aethiopica* proteome-wide prediction outputs to the experimentally acquired immunopeptidomics data of the clinical samples for which we had full HLA genotyping data (**Fig. 6C and Fig. 6D**). We predicted *L. aethiopica* MHC-binding peptides for 9 patients of experimental batch #3, and 4 patients of experimental batch #4. In total, NetMHCpan and NetMHCIIpan predicted 5.991.913 MHC-I and 14.607.989 MHC-II peptides for the clinical samples, respectively. For MHC-I, NetMHCpan correctly predicted 144 (71.6%) of the 201 epitopes observed to be MHC-presented in the immunopeptidomics experiments on clinical samples (**Fig. 6C**). Similarly, NetMHCIIpan predicted 138 (80.7%) of the 171 MHC-II epitopes observed in the immunopeptidomics experiments on clinical samples (**Fig. 6D**). Our findings indicate that, while immunopeptidomics only identifies the tip of the iceberg (likely the most abundant proteins), still ∼60-70% and ∼20-30% of these MHC-presented peptides were missed by the prediction tools in the *in vitro* and clinical experiments, respectively, despite the tools predicting millions of strong- and weak-binding peptides.

**Figure 6.**
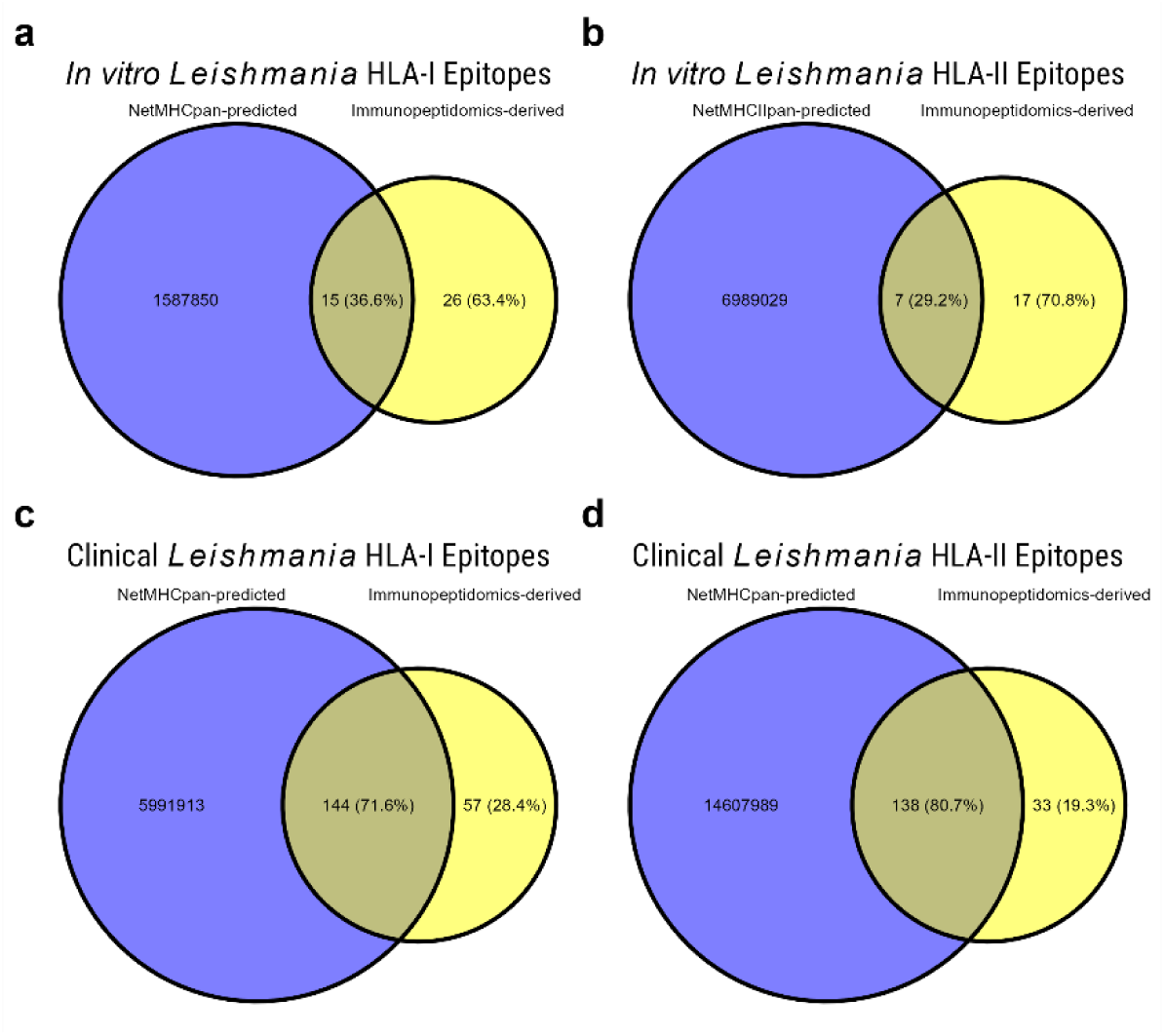
Overlap between experimentally observed peptides and in silico predicted peptides. Overlap between the L. aethiopica peptides experimentally observed using mass spectrometry-based immunopeptidomics, and in silico predicted L. aethiopica strong- and weak-binding peptides, using NetMHCpan and NetMHCIIpan for HLA-I and HLA-II, respectively. A) Overlap between experimentally observed (yellow) and predicted (blue) HLA-I peptides of the in vitro L. aethiopica-infected THP-1 cell line, B) Overlap between observed and predicted HLA-II peptides of the in vitro L. aethiopica-infected THP-1 cell line, C) Overlap between observed and predicted HLA-I peptides of the clinical samples, D) Overlap between observed and predicted HLA-II peptides of the clinical samples. Percentages shown are of the total experimentally observed L. aethiopica epitopes per HLA class.

## Discussion

We carried out the first immunopeptidome profiling of skin lesions from CL patients, and provide new insights in MHC-processed *Leishmania* proteins and novel vaccine targets which could not be identified with common prediction tools.

We successfully identified 333 MHC-I and 247 MHC-II-presented *L. aethiopica* epitopes derived from 398 *L. aethiopica* antigens in lesional biopsies from Ethiopian CL patients. According to IEDB, all the 580 *L. aethiopica* epitopes were novel and dissimilar to known *Leishmania* epitopes ^4^. We showed a significant degree of sharing between individuals (often with similar HLA genotypes) at the exact epitope level and also at the protein level, with several epitope-rich proteins. Combined with a high sequence identity between orthologous proteins across other *Leishmania* species for the top 10 antigens, we suggest those immunoprevalent proteins as promising vaccine antigens requiring further immunogenicity research. Moreover, using a cell line of *L. aethiopica*-infected THP-1 monocytes, we indicate that some of these epitopes and proteins might also be presented early on in infection.

Our analysis identified 51 antigens whose epitopes were presented by multiple, genetically diverse patients, suggesting they could be promising vaccine antigen candidates with broad population coverage. Among these, Elongation Factor 2 (EF-2) and Heat Shock Protein 70 (Hsp70) were the most prominent, showing shared presentation in five patients (18.5% of the study group). EF-2 was also the most epitope-rich antigen, suggestive of an immunoprevalent and/or immunodominant protein. It was, however, not detected in the early *in vitro* infection samples. This can either be the result of differential antigen processing *in* vitro versus *in vivo,* or a low abundance in the early *in vitro* infection and not meeting the limit of detection, or it can indicate restricted presentation in the later stages of the infection. Interestingly, EF-2, which is a GTP-binding translation elongation factor, has previously been described to be immunogenic in visceral leishmaniasis (VL) ^25–28^. In a first study by Kushawaha et al., a recombinant EF-2 protein (rEF-2) was used to stimulate PBMCs from cured and active VL patients, which eliciting a Th1-dominant immune response ^27^. In addition, they showed intradermal rEF-2 vaccination protected hamsters against a *L. donovani* challenge. Likewise, Joshi et al. also showed that *in silico* predicted and subsequently synthesized peptides of rEF-2 provided as intradermal vaccination induced a Th1 response and provided protection against *L. donovani* challenge in hamsters ^25^. Next, Agallou et al. pulsed dendritic cells with the N-terminal of EF-2, and showed Th1-dominant immune responses and protection against *L. infantum* challenge in mice ^26^. It is, however, important to note that the MHC-presented epitopes we identified are not in this N-terminal region. EF-2, together with Alpha Tubulin, were also shown to be immunogenic in a study by Probst et al. using expression cloning and T cell library screenings ^29^. As a follow-up to the Kushawaha et al. study, the authors showed that a combination of the recombinant Heat-shock Protein Hsp70 and recombinant EF-2 led to greater Th1 responses in VL patients and challenged animals than either alone ^28^. In line with their work, we also identified MHC-presented epitopes derived from the highly conserved multifunctional *Hsp70* protein of *L. aethiopica*, which were shared between individuals. *Leishmania Hsp70* was shown to be a potent adjuvant when combined with various *Leishmania* antigens in experimental *Leishmania donovani* models for VL ^30,31^. Both Hsp70 and Alpha Tubulin were proposed as new diagnostic target antigens for VL based on expression cloning and immunoblotting with sera of Indian VL patients ^32^. The DNA vaccine HISA70, which combines Hsp70, Histone H2A, other nucleosomal histones, and the A2 protein, also conferred protection against *L. major* infection in mice ^33^. Histone H2A is the top epitope-rich antigen in our study when including our early infection *in vitro* samples. It is a confirmed B cell antigen for canine leishmaniasis, and has been included in the veterinary vaccine LetiFend ^34^. Histone H2A, together with other nucleosomal histones, has also been shown to confer protection against murine CL ^35,36^. Antigens used in prior human vaccine trials, that were commonly based on immunodominance in small animal models, include thiol-specific oxidant (TSA), Cysteine protease A and B (CPA and CPB, respectively), Hydrophilic acylated surface protein B (HASPB), Kinetoplastid Membrane Protein 11 (KMP-11), *L. major* stress-inducible protein 1 (lmSTI1), Amastigote stage-specific protein (A2), recombinant *Leishmania* elongation factor 1α (P74), Nucleoside Hydrolase (NH36), and Sterol 24-c-methyltranferase (SMT). Remarkably, while all these antigens showed immunogenicity in the early phase trials, we only identified one epitope presented of the *L. aethiopica* orthologous protein of lmSTI1, and none of the other antigens were observed to be presented, although immunopeptidomics detection limits may have prevented us from detecting these. This suggests a combination of known vaccine candidate antigens with our new observed MHC-presented antigens could potentially form more promising targets for a human vaccine candidate against CL, perhaps even VL, although follow-up research is needed on early availability/presentation and the functional phenotype/efficacy of induced T cells to these antigens in a human infection.

Peptide-MHC binding prediction tools are widely used for *Leishmania* antigen discovery ^37,38^. Here, we observed that while the popular prediction tools NetMHCpan and NetMHCIIpan correctly predict around a third of our experimentally-observed epitopes in the *in vitro* experiments, and around 70-80% in the clinical samples, they still missed a significant proportion of likely high abundant processed proteins and predicted millions of potential peptides that may not be naturally presented. It should be noted that the 70–80% observed in clinical samples likely represents an overestimation as well, as the comparison was performed by pooling all peptides predicted across all HLA alleles with all peptides identified by immunopeptidomics. Mass spectrometry-based immunopeptidomics does not inform us of which peptide binds to which HLA allele, and thus, using the pooled approach, a peptide predicted for one allele (e.g. HLA-A*03:01) may be counted as a match even if the experimentally observed peptide only binds HLA-A*02:01 in biological reality. Given these data, for *Leishmania* reverse vaccinology or vaccine discovery purposes, prediction-only pipelines risk capturing false positives while missing real MHC-presented peptides that could be critical, and careful considerations should be taken when *in silico* peptide-MHC binding prediction tools are used for these purposes. The low predictive performance can be partially explained by a number of reasons. Firstly, although the mass spectrometry-based immunopeptidomics epitope identification rate has drastically improved, several issues remain that may form a bottleneck in identifying all MHC-presented epitopes, such as the extensive search space due to the absence of strict digestion rules ^39,40^. Secondly, the *in silico* peptide-MHC binding prediction tools NetMHCpan and NetMHCIIpan do not take into account parasite protein abundance, a factor that has been shown to increase predictive performance for SARS-CoV-2 epitope prediction once taken into account ^41^. Similarly, they do not take into account potential parasite antigen machinery escape mechanisms that might heavily affect protein availability, abundance, and cleavage ^42^. As there is a multitude of prioritization strategies, future research should assess whether any prioritization strategy, such as those working by subtractive proteomics or molecular docking, will reduce the peptides to screen experimentally substantially while preserving the correct MHC-presented peptides ^7,43^.

While multiple studies report MHC downregulation *in vitro* during *Leishmania* infection, this has not been shown in patients ^44,45^. Although we cannot infer whether MHC antigen presentation was reduced, we still identified many MHC-presented epitopes, showing an active and detectable MHC presentation from early to late infection despite potential downregulation. To account for the inclusion of patients with relatively late stages of disease presentation, we included *in vitro* cell lines at early stages of parasite infection to infer whether certain antigens were already presented as early as 5h, 24h, 72h post-infection.

A limitation of our study is that patient outcome data was missing for most of the patients, which restricted our ability to assess whether antigen MHC presentation was linked with clinical outcomes. Future work should ideally include complete patient disease outcome data and longitudinal sampling to monitor antigen presentation dynamics through the disease or to map the impact of treatment on antigen presentation. Recently, a controlled human infection model (CHIM) was developed for CL caused by *L. major*, which may enable us to more easily include such factors, allowing for a more precise assessment of antigen dynamics in a controlled setting, and will significantly expedite antigen discovery and subsequent vaccine development and evaluation ^46^. Another limitation of this study is that we did not have healthy endemic control samples to verify whether the MHC-presented peptides observed were truly unique to CL. In addition, in this study, we used the online available *L. aethiopica* reference proteome instead of personalized clinical isolate proteomes. While personalized clinical isolate proteomes are costly and hard to get, they can increase the number of confidently identified *Leishmania* peptides. Despite this, with a total of 650 *L. aethiopica* peptides, we were still able to discover 28-fold more peptides than the currently known 23 *Leishmania* peptides MHC-presented *in vitro*, and roughly 3-fold more than the 228 *Leishmania* T cell epitopes currently listed on the IEDB ^4^. In addition to the above mentioned problem, the *L. aethiopica* proteome is poorly annotated, thus limiting the information on protein function for many of the proteins as evidenced by the many hypothetical proteins that require follow-up research on their function/importance. This mostly affects the Gene Ontology enrichment analysis which showed that there was an overrepresentation of antigens involved in ribosomal function. However, this is in line with observations that defective ribosomal products are a major source of antigenic peptides for MHC-I ^47^.

In conclusion, our findings open up the way for a more in depth characterization of the *Leishmania* protective T cell response, or the T cell receptor specificity for *Leishmania* epitopes, an avenue that has until now been scarcely explored for *Leishmania*, but has already provided important insight on antigen selection and disease determinants for *Mycobacterium tuberculosis* vaccine development ^48^. We believe the MHC-presented *L. aethiopica* epitopes and antigens that are most shared across patients represent valuable vaccine candidates. In addition, these targets enable further exploration of peptide-MHC-TCR interactions, and will allow T cell receptor specificity screening, which can in turn be leveraged to better define the protective T cell response.

## Data Availability

All data produced in the present work are contained in the manuscript or available upon reasonable request to the authors

## Author contributions

N.d.V., T.T.P., P.M., B.C., K.L. and W.A. designed and conceptualised the study. E.P. and L.T. performed the mass spectrometry experiments. N.d.V., L.L. and K.B. performed the data analyses. N.d.V. created all figures. N.d.V., T.T.P., and I.M. performed the cell culturing. N.d.V. and K.R. performed DNA extractions and HLA genotyping. Sample isolation and clinical work was performed by T.M., M.W., S.H., F.B., S.v.H., and F.T. Sample handling and storage was performed by M.K. Clinical study coordination was done by Y.A., M.S.A., M.W, S.G.A, M.A.D, S.v.H., J.v.G and W.A. N.d.V. wrote the manuscript. N.d.V., E.P., L.L., T.T.P., M.A.D., B.C., P.M., S.v.H., J.v.G., K.L., G.B., and W.A. reviewed and edited the manuscript. B.C., K.L., P.M., and W.A. supervised the manuscript preparation. N.d.V., B.C., G.B., J.v.G., S.v.H., K.L., and W.A. acquired the funding for the study. All authors have read and agreed to the published version of the manuscript.

## Funding statement

This work was supported by the Dioraphte Foundation (the Netherlands) under the skin NTD programme, the Department of Economy, Science and Innovation of the Flemish Government under ITM’s Pump Prime Project programme, the Belgian Directorate General for Development Cooperation under the ITM-DGD framework agreement IV, a research grant of the University of Antwerp Research Fund (BOF) [FFB220027 to B.C. and N.d.V.], an through a fellowship (1S71721N) to N.d.V. by the Research Foundation Flanders (FWO). The funders had no role in study design, data collection and analysis, decision to publish, or preparation of the manuscript.

## Competing interests

W.A., K.L, T.T.P, M.S.A. and N.d.V have filed a patent (EP25207271) for vaccine candidates based on the results of this manuscript. The institutions involved in this patent are the University of Antwerp, the Institute of Tropical Medicine Antwerp, and the University of Gondar.

## Acknowledgements

Our gratitude goes to all participants involved in this study, and all the Leishmaniasis Treatment and Research Center and Boru Meda Hospital staff members who were actively involved in the care of these participants. The authors also thank the Drugs for Neglected Disease Initiative and the University of Gondar for supporting the Leishmaniasis Research and Treatment Center. Finally, N.d.V. wants to thank his dearly beloved late fiancée Sofie Van de Poel for her eternal love and support.

