## Supplementary figures and images for "Immunopeptidomics of cutaneous leishmaniasis patients reveals the natural antigenic landscape"

### Supplemental Figure 1

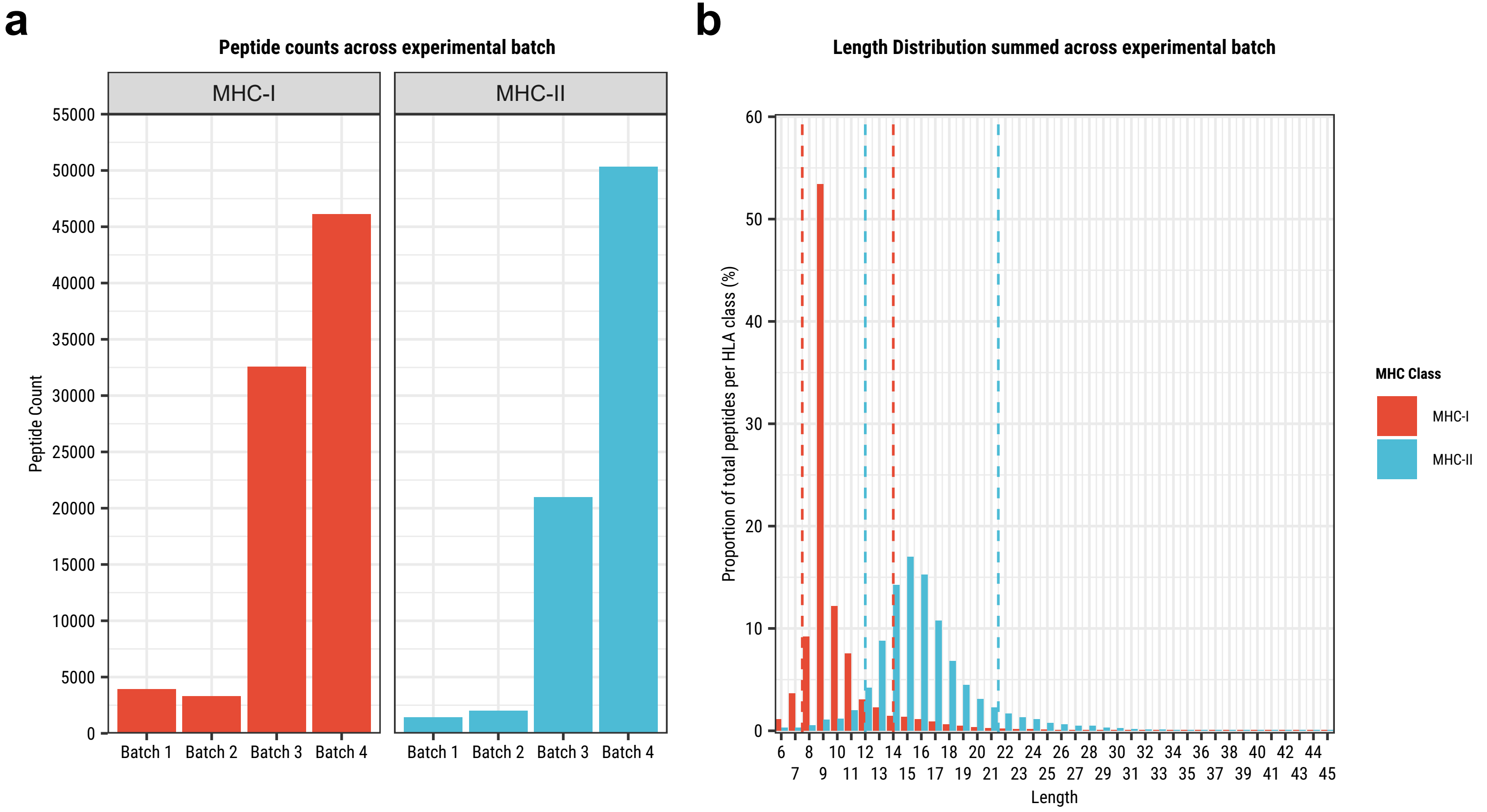

### Supplemental Figure 2

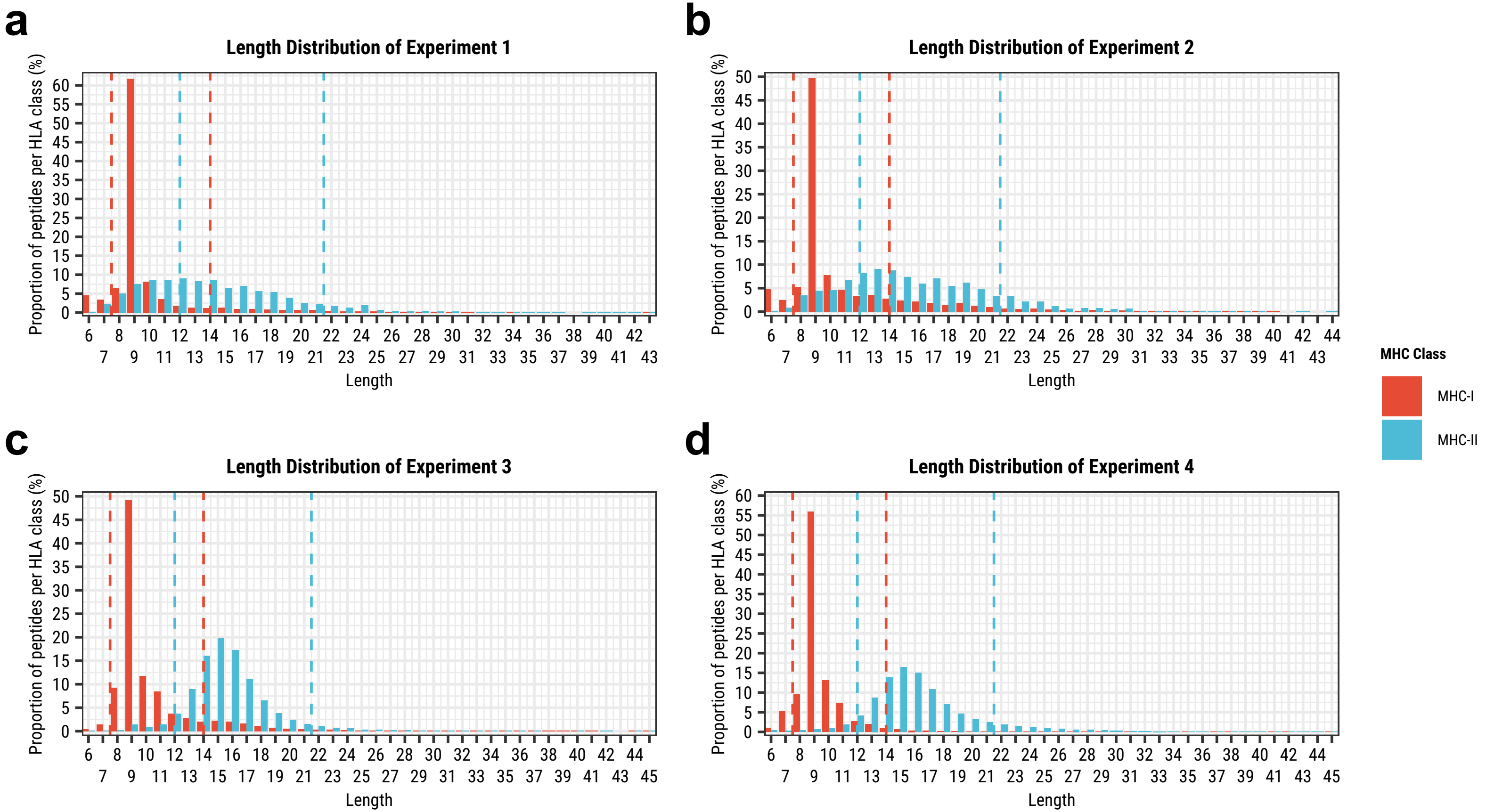

### Supplemental Figure 3

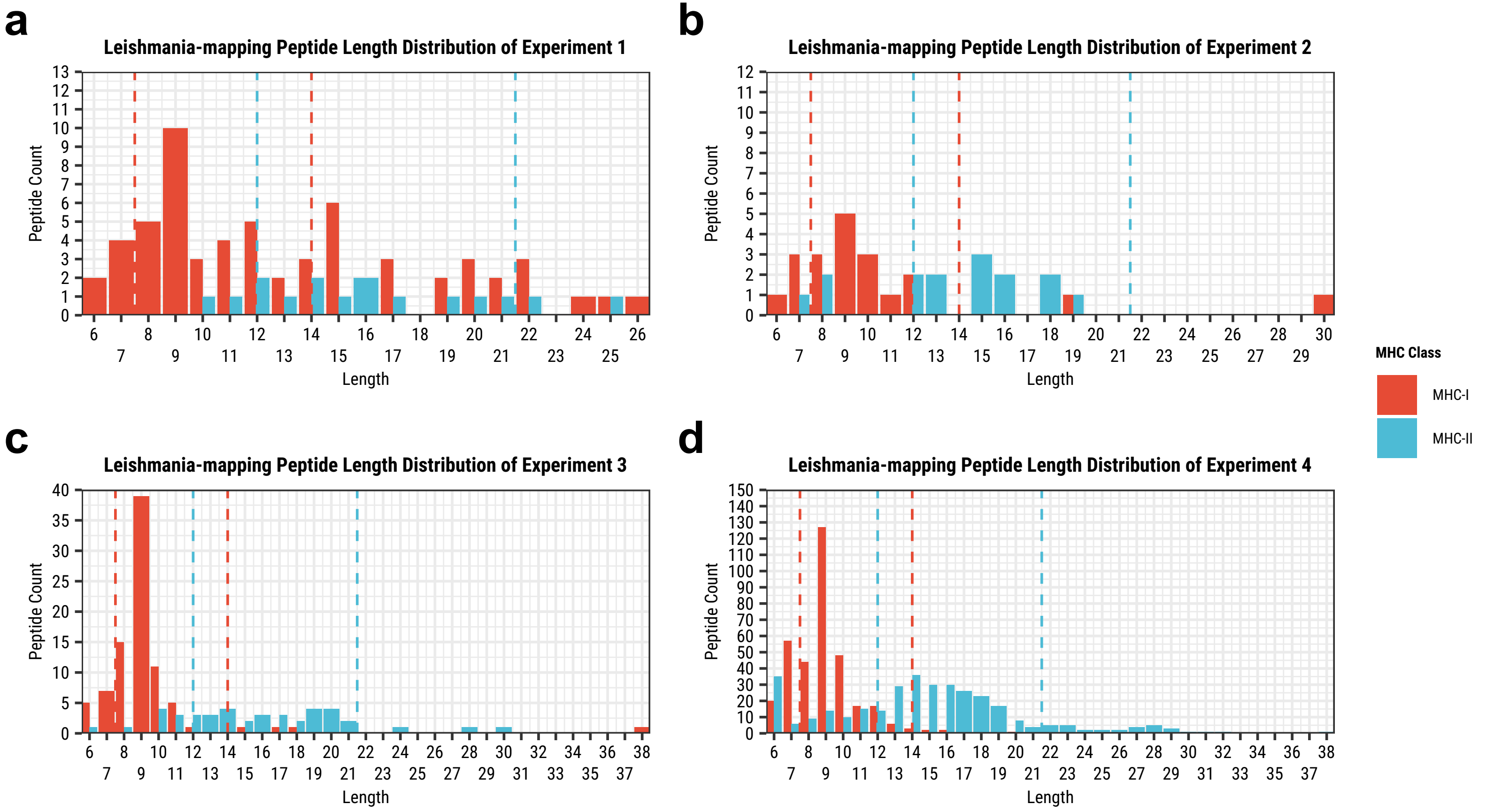

### Supplemental Figure 4

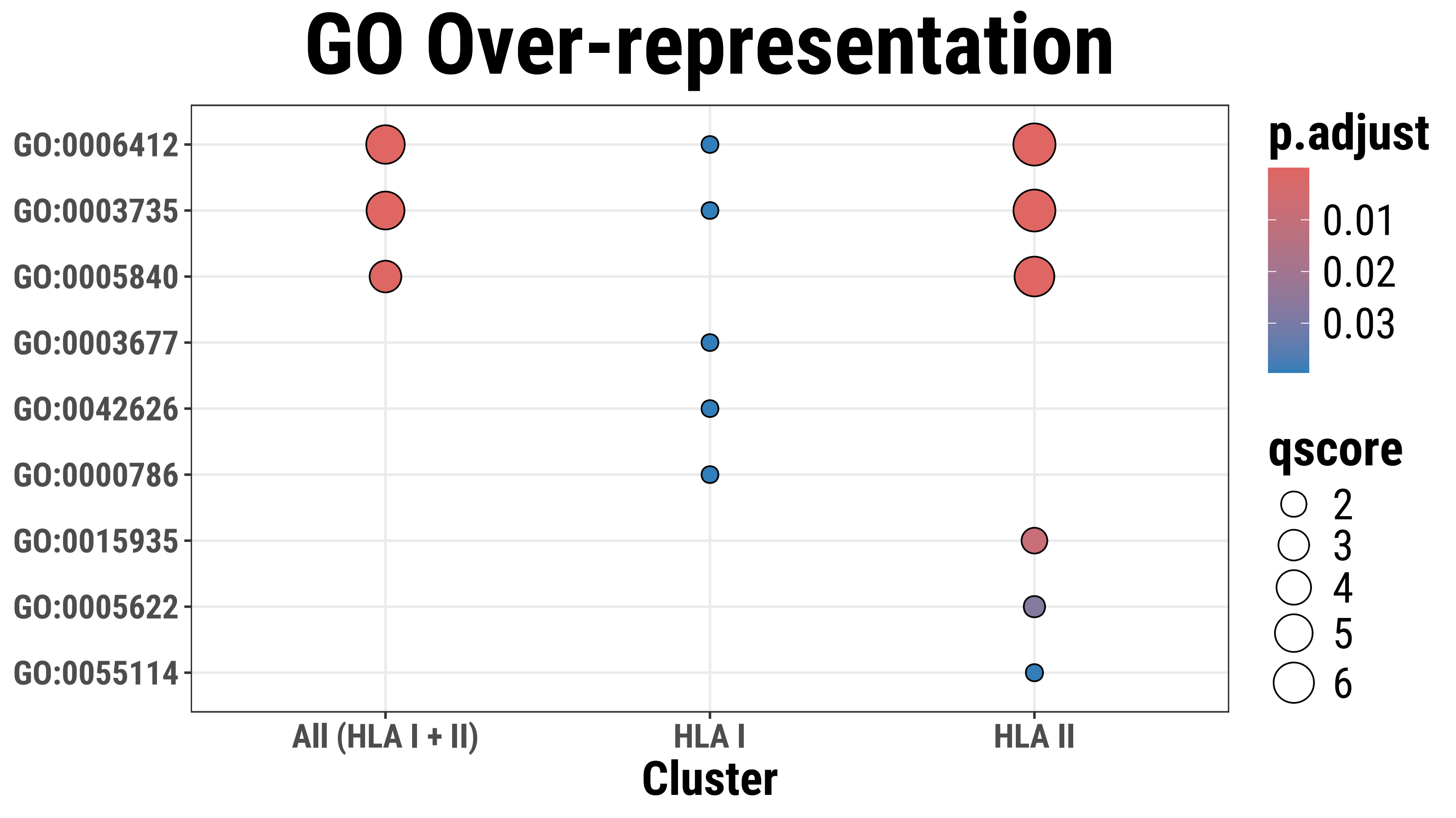
